# Human-centered design can optimize operational details of an implementation strategy: Experiences from a peer-based hypertension medication delivery program in western Kenya

**DOI:** 10.64898/2026.10.29.26366812

**Authors:** Mercy Korir, Hamza Ahmed, Benson Njuguna, Jessica Gjonaj, Kibet Kangogo, Jeremiah Laktabai, Caroline Watiri Maina, Imran Manji, Beryl Masinde, Sonak Pastakia, Dan N. Tran, Juddy Wachira, Rajesh Vedanthan

## Abstract

**Background:** Human-centered design (HCD) is a co-creation approach that is increasingly being utilized by implementation science researchers to engage key stakeholders in developing contextually appropriate health interventions and implementation strategies. The exact role of HCD in implementation research, however, remains uncertain. We describe the process, outcome, and experiences of using HCD to develop the operational details of an implementation strategy based on a pre-existing scaffolding that consisted of combining community-based peer medication delivery with health information technology (HIT) to improve medication adherence among hypertension patients in rural western Kenya.

**Methods:** A design team of 22 members was formed representing key stakeholders: hypertension patients, healthcare workers, pharmacists, peers, HIT personnel, policy/program leaders, and implementation researchers. We conducted six meetings utilizing established HCD principles and processes (inspiration, brainstorming, conceptualization, creation, iteration, and finalization), aimed at developing a final implementation strategy with full operational details. Quantitative assessment of acceptability and appropriateness was conducted among healthcare workers, patients, and peers, using the Acceptability of Intervention Measure and Intervention Appropriateness Measure.

**Results:** The design team developed key operational details across five domains: peer recruitment, training and monitoring; HIT; medication delivery; medication payment; and stakeholder identification. Acceptability and appropriateness scores (maximum of 20) were favorable among all participants, but mean scores were notably higher among patients with hypertension (19.3, 19.2) and peers (19.2, 19.2) compared to healthcare workers (17.7, 17.3). Specific recommendations were made to improve the acceptability and appropriateness of the strategy. These findings were subsequently used to develop a final iteration of the implementation strategy.

**Conclusion:** HCD was feasible and useful in optimizing the operational details of our implementation strategy, aligning it with the healthcare goals and needs of the community. It allowed end-users to co-create with researchers, ultimately yielding high pre-implementation acceptability and appropriateness, which we anticipate will ultimately enhance implementation success.

## BACKGROUND

Despite an increasing burden of hypertension in low- and middle-income countries (LMIC), blood pressure control rates in LMICs are low.^1^ These low control rates are concerning given the adverse cardiovascular effects and death resulting from complications of uncontrolled elevated blood pressure, with elevated systolic blood pressure ranking as a top contributor to disability-adjusted life years in LMICs.^1^ In Kenya, hypertension prevalence is estimated at 24% and control rates are less than 20%.^1^ Medication nonadherence in Kenya is a significant cause for these low control rates, with approximately 71% of participants with hypertension and/or diabetes reporting sub-optimal medication adherence in a recent cross-sectional study from western Kenya.^2^ Significant barriers to adherence, such as transportation costs that influence medication access and possession, as well as inadequate psychosocial support, need to be addressed to improve health status.^2^

Novel interventions leveraging community health workers and peer networks have been effective previously in improving diabetes and HIV control in Africa, and have the potential to address the aforementioned barriers to ultimately improve medication adherence and blood pressure control rates.^3,4^ In addition, health information technology (HIT) and digital health interventions incorporating reminders, decision support, patient tracking, and electronic medical records have been shown to be effective at improving medication adherence.^5^ However, the combination of peer coaching, community-based medication delivery, and HIT support to improve medication adherence and clinical outcomes for people living with hypertension has not been well studied. Thus, our team plans to implement and evaluate an implementation strategy, known as the Peers and Technology for Access, Adherence, Accountability and Analytics (PT4A), to address this gap.

In order to create successful interventions and implementation strategies that will lead to high end-user satisfaction and engagement, the desires and interests of the end-users must be taken into account, especially for interventions and implementation strategies involving multiple community stakeholders.^6^ The human-centered design (HCD) approach is gaining popularity and has shown success in implementation research studies in low- and middle-income countries.^7,8^ The HCD approach involves deep discussion and exploration with end-users utilizing interactive workshops and interviews to generate tailor-made solutions that fit their needs and are feasible and appropriate for society. By involving stakeholders and community members in the design process, interventions and implementation strategies tend to be more appropriate and acceptable for the end-users for whom they are developed, leading to better outcomes.^9,10^

In prior published studies of the HCD process, most interventions and implementation strategies were already designed or in place, and the HCD process was used to refine these interventions and provide researchers with insights into public perceptions of their interventions. For example, the BIGPIC study in western Kenya applied the HCD process to refine a hypertension and diabetes care delivery model combining microfinance and group medical visits that had been previously developed and piloted.^7,11^ HCD has been used in a similar manner for a number of other interventions in several African countries.^12–16^ However, it has not yet been described how to leverage HCD to specify operational details for a planned implementation strategy. Therefore, we aimed to use the HCD process to generate operational details for the PT4A strategy.

## METHODS

### Study setting

This study was conducted in western Kenya within the context of the Academic Model Providing Access to Healthcare (AMPATH), chronic disease management (CDM) program. AMPATH is an academic partnership between Moi University College of Health Sciences (MUCHS), Moi Teaching and Referral Hospital (MTRH), and a consortium of North American universities.^17^ AMPATH established a CDM program in 2011 that includes several components such as task redistribution, clinical decision support using HIT, consistent and secure medication supply, linkage and retention activities, community stakeholder engagement, and social support including MF groups.^9,18–21^ CDM has enrolled more than 60,000 patients in over 150 health facilities in its catchment area in western Kenya.^22^ Our study took place in three of the AMPATH catchment areas: Webuye in Bungoma County, Kitale in Trans-Nzoia County and Turbo in Uasin Gishu County.

### Core components of the implementation strategy

The proposed implementation strategy has three core components: peer coaching, community-based medication delivery and HIT (**Figure 1**). Peers are lay individuals with shared disease experience that can be leveraged to provide peer coaching consisting of practical, social, emotional, and motivational assistance to patients.^23^ Peer coaching will address individual-level adherence barriers, provide psychosocial support, and enhance patient activation. This peer coaching will be offered during the door-to-door community-based delivery of medicines to patients with hypertension at convenient locations and time collaboratively decided by patients and peers. The HIT component will be aligned with AMPATH informatics infrastructure to provide decision support and tailored adherence counseling.^24,25^ This component will also facilitate tele-consultation between patients and clinicians and will be used for tracking and accountability by providing prompts to peers when refills are near due.

**Figure 1.**
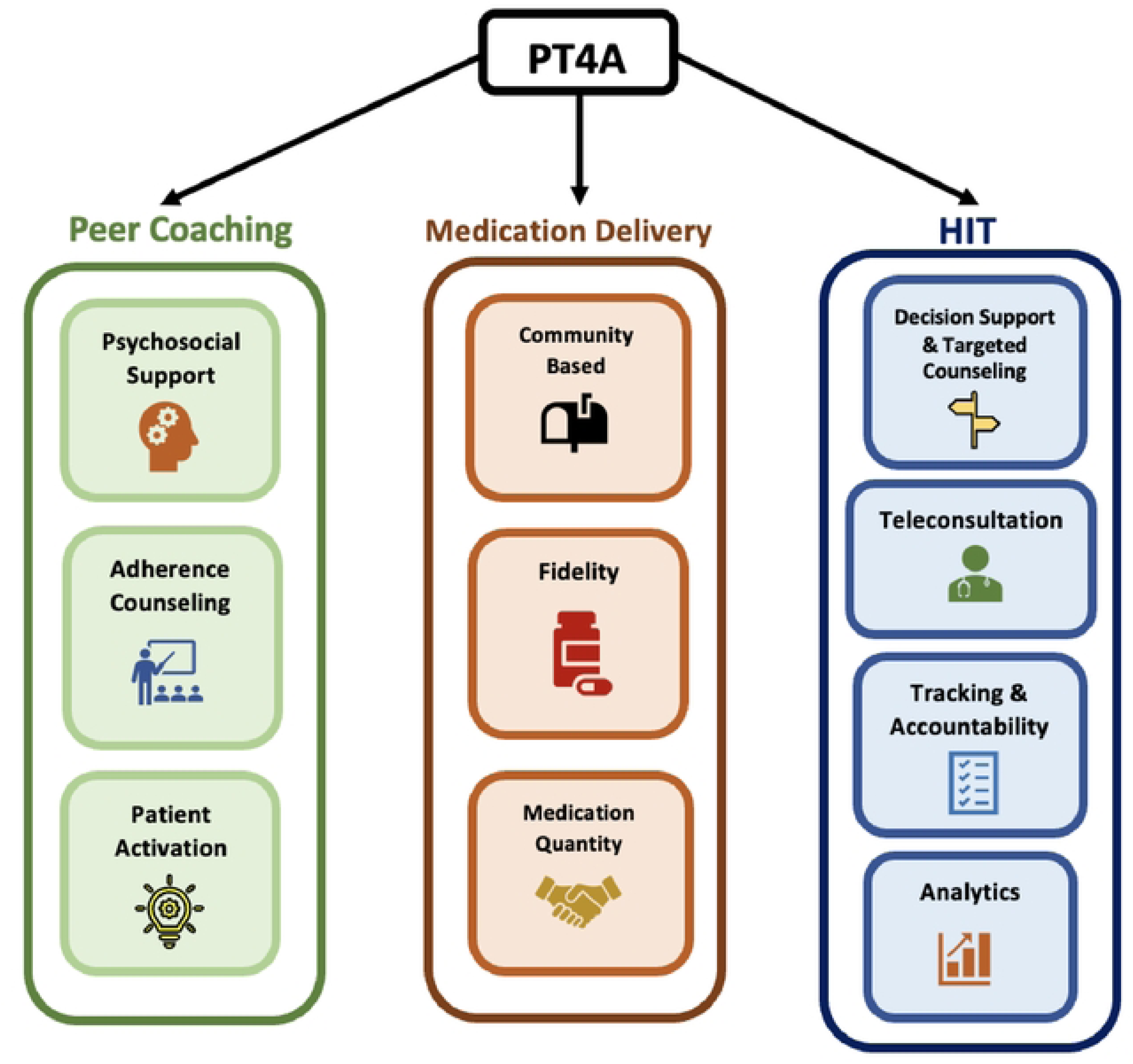
Core components of the implementation strategy. Diagram illustrating peer delivery and HIT (health information technology) components of the PT4A implementation strategy as well as how the implementation strategy functions to improve medication delivery and adherence.

### Design team facilitator training

The study team received a two-month online training on ‘Introduction to Human-Centered Design’ from Acumen Academy, which is supported by IDEO.^26^ Each module had activities and discussions that were required to be completed in groups and updated before moving to the next module. The study team held weekly online discussions that lasted approximately one hour each. In addition to online meetings, the group created online boards that were accessible to each team member to share their perspectives for further discussion. Every member of our study team completed all four modules of the IDEO training course.

### Design team formation

The design team was comprised of 22 members representing the targeted end users: physician (1), clinical officers (2), nurse (1), pharmacists (2), peer navigators (3), patients with hypertension (2 male, 2 female), HIT personnel (3), national health insurance fund representatives (2), and research team members (4). Patients were randomly selected during their routine clinic visits at the various facilities, one from Kitale, one from Turbo, and two from Webuye. After approval from hospital administration had been obtained, healthcare workers were selected from those who were currently engaged in patient care in the CDM clinics of the various facilities within the study sites. HIT personnel were selected from those who had experience with the AMPATH medical records system. Peers had to be active in their peer roles within various programs in AMPATH. Representatives from the national health insurance fund (the Kenyan national public health insurance scheme), were also included.

Once identified, the research team briefed them on the HCD process, the number of expected sessions, and the duration of each session. All participants who were willing and able to attend all the sessions were invited. They were from different health facilities across the three study sites. For logistical purposes, the design team members from all the study sites were convened in one central geographical location which enabled in-person participation of the design team members.

### Design process

We conducted a total of six design team meetings (**Figure 2**): inspiration (meeting #1), brainstorming (#2), conceptualization (#3), creation (#4), iteration (#5), and finalization (#6). This paper describes the first five meetings and presents the results of acceptability and appropriateness testing conducted after meeting #4, focusing on how we leveraged HCD to optimize the operational details of the implementation strategy in preparation for the pilot study. The pilot study implementation and feasibility testing, as well as the outcomes of meeting #6 (finalization of the implementation strategy), will be described and reported separately in an independent manuscript.

**Figure 2.**
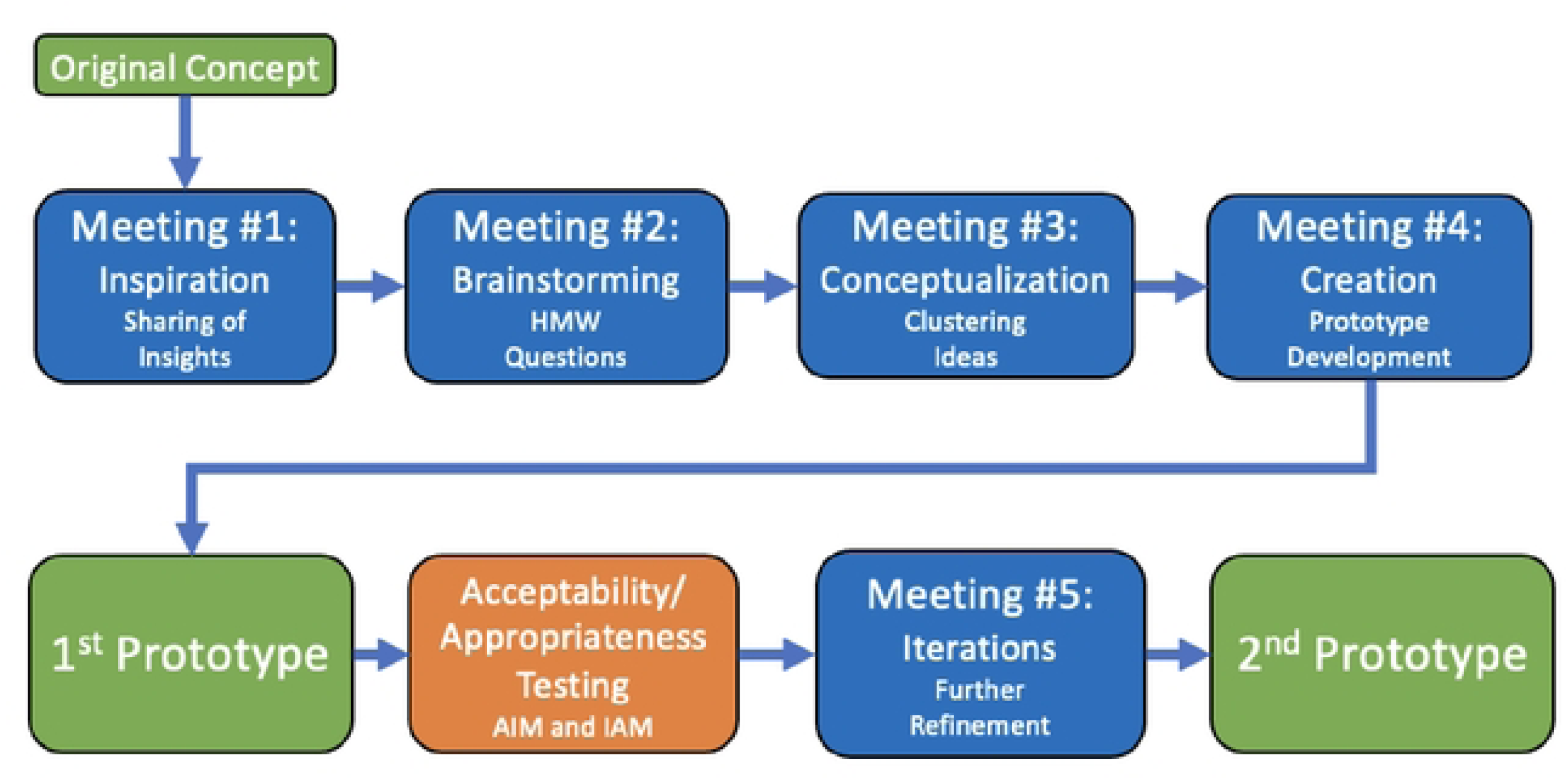
Design process. Illustration describing the design process of the PT4A strategy including the various meetings that occurred, testing that was done, and prototypes created. Blue boxes indicate meetings that occurred, green boxes indicate prototypes, and orange indicates a testing period. *\*HMW = “How MightWe”, AIM = Acceptability of Intervention Measure, IAM = Intervention Appropriateness Measure*

The first four meetings were weekly meetings, each approximately five hours long, anchored on the core components of the implementation strategy as described above. The prototype generated after meeting #4 underwent acceptability and appropriateness testing. This testing process and analysis took approximately three months, after which meeting #5 was conducted. Each meeting’s agenda was developed and led by two facilitators (JA, EN) who had received training and had prior experience in HCD implementation. All design team meetings were collaborative and interactive.

### Design meetings

#### Meeting 1-Inspiration

The goal of this meeting was to introduce the team to HCD, present baseline findings and identify key refinement areas. To better understand HCD, team members were paired up and asked to identify and discuss the challenges they faced during their commute to the workshop that morning, and to explore possible solutions. This exercise laid the groundwork for a discussion on the meaning and application of HCD.

The study team then presented the findings from the baseline contextual assessment, which aimed to identify factors that could influence the operational details of the implementation strategy.^27^ The findings highlighted several perceived benefits of the planned strategy, including improved hypertension management, enhanced psychosocial support, better community-based follow-up for patients, and reduced healthcare costs. The assessment also identified areas requiring further refinement, including the ideal qualifications of peers, medication payment processes, delivery procedures, and specifications for the HIT component. Following the presentation, team members reviewed the findings and shared their personal experiences and insights on aspects of the implementation strategy that needed further development. Each member documented their reflections on Post-it notes, which were then organized on a board to guide discussion and prioritization.

#### Meeting 2-Brainstorming

The goal of this meeting was to translate insight statements into ‘How Might We’ questions to stimulate creative thinking and to guide generation of potential solutions. The team used an iterative process to refine the initial insight statements into final core statements that were relevant to the implementation strategy. These final core insight statements were then reframed as ‘how might we’ questions which created an exploratory space for identifying and discussing possible solutions.

#### Meeting 3-Conceptualizing

The goal of this meeting was to evaluate and refine the proposed solutions in order to develop implementation components that were acceptable, feasible and aligned with the implementation strategy. Members reviewed solutions generated in meeting 2 and assessed each for anticipated feasibility, potential advantages, and potential disadvantages. During the meeting, the team continued to generate additional ideas and solutions while also adding elements to existing ones. Any ideas that were deemed unfeasible or not aligned with the core components of the implementation strategies were discarded. Similar concepts were grouped together. This process was cyclical and iterative, continuing until members reached agreement on final solutions with feasible operational components.

#### Meeting 4-Creation

The goal of this meeting was to synthesize the refined concepts from previous meetings into a visual workflow and to prepare the first prototype of the implementation strategy for acceptability and appropriateness testing. Research team members presented the detailed concepts of the proposed solutions, while other members provided feedback, either agreeing with the proposed recommendations or suggesting further modifications based on their potential to strengthen the implementation strategy.

During the meeting, the study informatics team also presented the proposed structure of the HIT tool, developed based on input from Meeting 3. Design team members reviewed the proposed workflow and provided feedback on the flow structure, required fields, and overall usability of the tool. By the end of the meeting, recommendations across all domains had been incorporated into a visual depiction of the first prototype implementation strategy, which was then ready for acceptability and appropriateness testing.

### Acceptability and Appropriateness testing

The prototype implementation strategy was then tested for acceptability and appropriateness based on an adapted version of the psychometrically validated and reliable Acceptability of Intervention Measure (AIM) and Intervention Appropriateness Measure (IAM).^27^ Acceptability is a measure of how innovation is perceived as agreeable or satisfactory whereas appropriateness measures its perceived relevance and compatibility.

A total of 39 participants were enrolled across all study sites: 15 patients with hypertension, 15 clinicians, 6 peer providers, and 3 pharmacists. Patients were recruited through random sampling during routine clinic visits. Clinicians, pharmacists, and peer providers were required to be actively involved in the clinical management of hypertensive patients, while all participants were required to have had no prior involvement in the HCD process. Written informed consent was obtained from all participants prior to enrollment.

Research assistants described the intervention in detail while displaying the visual depiction created in meeting #4 for better understanding. Thereafter, each participant rated the acceptability and appropriateness of the intervention. Acceptability and appropriateness scores were summed separately. Each of these domains has four items with a 5-point response scale ranging from ‘completely disagree’ (a score of 1) to ‘completely agree’ (a score of 5). Total scores for each domain thus range from 4-20.

#### Meeting 5-Iteration

This meeting followed the acceptability and appropriateness testing, and the goal was to refine the prototype based on feedback from the testing. The study team presented their findings, focusing on areas with low scores on either acceptability or appropriateness measures. The team then formed ‘how might we’ questions to prompt further brainstorming. For example, if group ‘x’ scored low on the overall acceptability measure, the HMW question was framed as “How might we make the implementation more acceptable among group ‘x’?” The design team then brainstormed possible solutions. At the end of this meeting, we had the second prototype implementation strategy for pilot implementation.

### Ethical considerations

This study obtained ethical approval from the Institutional Research and Ethics Committee at Moi University, College of Health Sciences in Eldoret, and the Institutional Review Board at NYU Grossman School of Medicine. ClinicalTrials.gov identifier (NCT number) is NCT05051124. All participants provided written informed consent before enrollment in this study.

## RESULTS

### Design meetings 1-4

In meeting 1, the deliberation process resulted in the organization of team members’ inputs into five operational domains that addressed the key factors identified during the baseline contextual assessment. These domains were peer recruitment, training, and monitoring; HIT; medication delivery; medication payment; and stakeholder identification (**Table 1**). These domains were further developed through reframing and solution brainstorming during meeting 2 and 3. In meeting 4, all recommendations were synthesized, and operational details for different domains were finalized, creating the 1^st^ prototype (**Table 1**). These refinements built on the originally conceptualized strategy by adding more specific details to improve feasibility, acceptability, and responsiveness to end users’ needs.

**Table 1:**
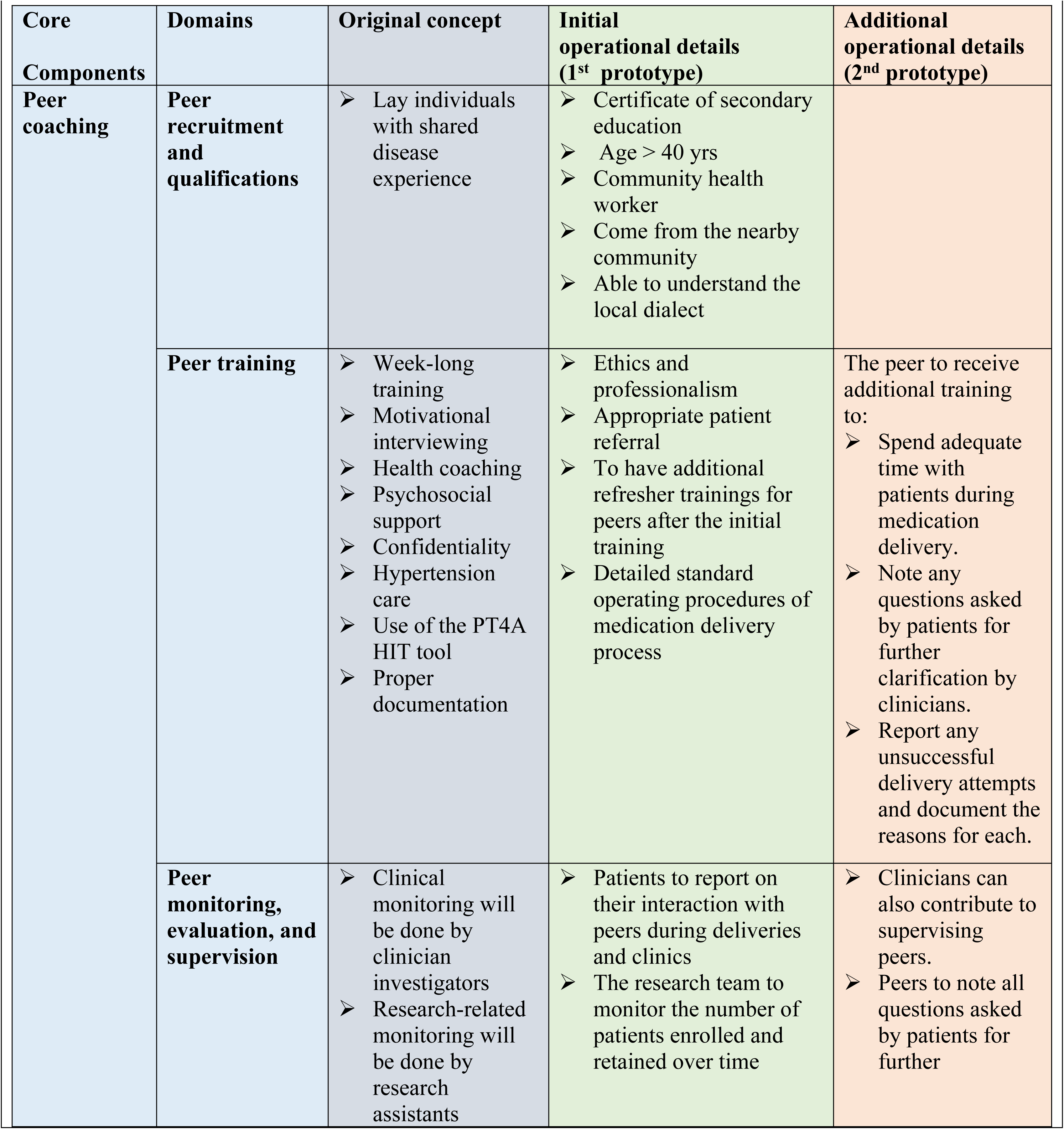
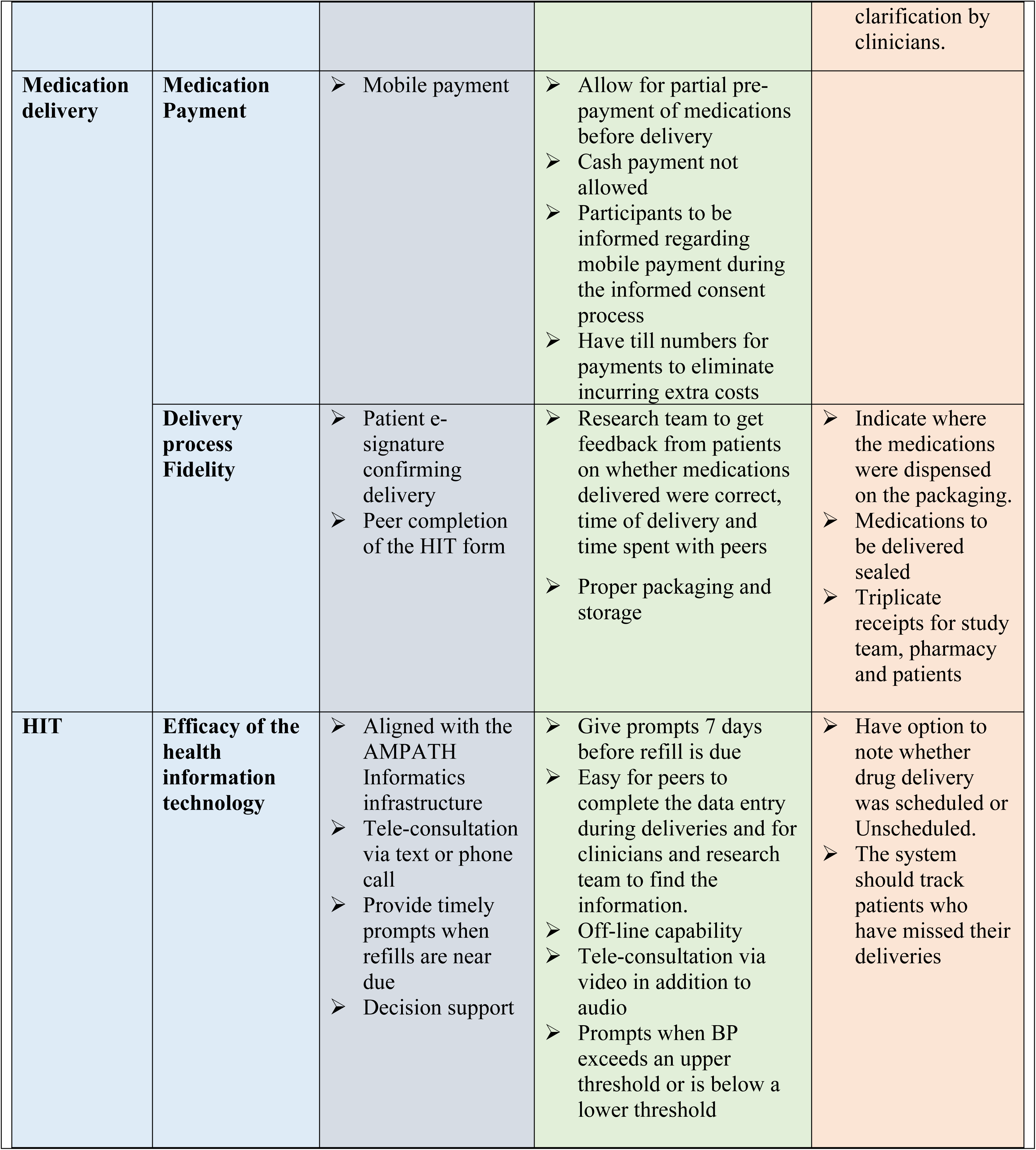
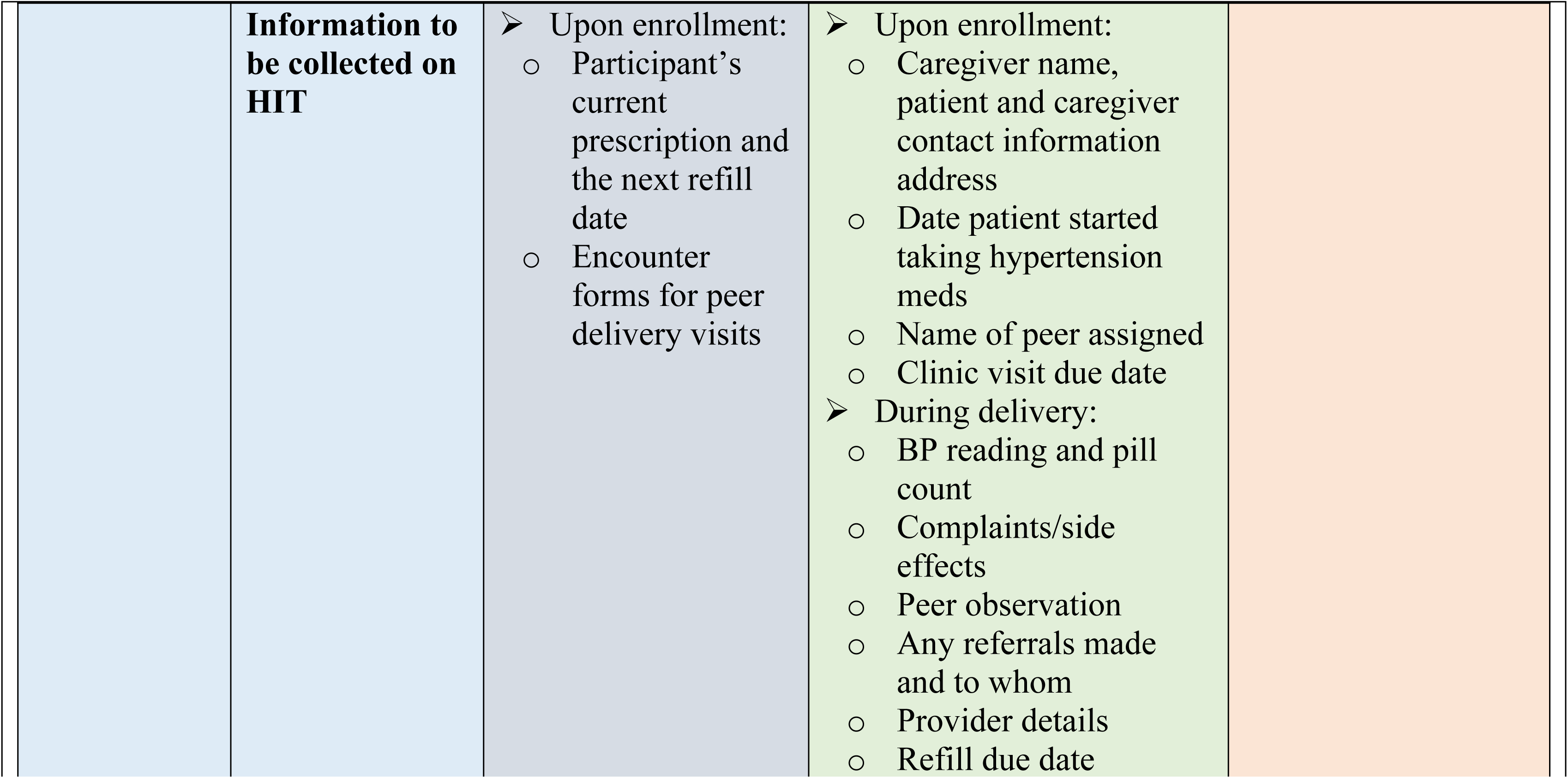
Implementation details added through HCD.

| Core Components | Domains | Original concept | Initial operational details (1<sup>st</sup> prototype) | Additional operational details (2<sup>nd</sup> prototype) |
|---|---|---|---|---|
| **Peer coaching** | **Peer recruitment and qualifications** | ➢ Lay individuals with shared disease experience | ➢ Certificate of secondary education<br>➢ Age > 40 yrs<br>➢ Community health worker<br>➢ Come from the nearby community<br>➢ Able to understand the local dialect |  |
|  | **Peer training** | ➢ Week-long training<br>➢ Motivational interviewing<br>➢ Health coaching<br>➢ Psychosocial support<br>➢ Confidentiality<br>➢ Hypertension care<br>➢ Use of the PT4A HIT tool<br>➢ Proper documentation | ➢ Ethics and professionalism<br>➢ Appropriate patient referral<br>➢ To have additional refresher trainings for peers after the initial training<br>➢ Detailed standard operating procedures of medication delivery process | The peer to receive additional training to:<br>➢ Spend adequate time with patients during medication delivery.<br>➢ Note any questions asked by patients for further clarification by clinicians.<br>➢ Report any unsuccessful delivery attempts and document the reasons for each. |
|  | **Peer monitoring, evaluation, and supervision** | ➢ Clinical monitoring will be done by clinician investigators<br>➢ Research-related monitoring will be done by research assistants | ➢ Patients to report on their interaction with peers during deliveries and clinics<br>➢ The research team to monitor the number of patients enrolled and retained over time | ➢ Clinicians can also contribute to supervising peers.<br>➢ Peers to note all questions asked by patients for further clarification by clinicians. |
| **Medication delivery** | **Medication Payment** | ➢ Mobile payment | ➢ Allow for partial pre-payment of medications before delivery<br>➢ Cash payment not allowed<br>➢ Participants to be informed regarding mobile payment during the informed consent process<br>➢ Have till numbers for payments to eliminate incurring extra costs |  |
|  | **Delivery process Fidelity** | ➢ Patient e-signature confirming delivery<br>➢ Peer completion of the HIT form | ➢ Research team to get feedback from patients on whether medications delivered were correct, time of delivery and time spent with peers<br>➢ Proper packaging and storage | ➢ Indicate where the medications were dispensed on the packaging.<br>➢ Medications to be delivered sealed<br>➢ Triplicate receipts for study team, pharmacy and patients |
| **HIT** | **Efficacy of the health information technology** | ➢ Aligned with the AMPATH Informatics infrastructure<br>➢ Tele-consultation via text or phone call<br>➢ Provide timely prompts when refills are near due<br>➢ Decision support | ➢ Give prompts 7 days before refill is due<br>➢ Easy for peers to complete the data entry during deliveries and for clinicians and research team to find the information.<br>➢ Off-line capability<br>➢ Tele-consultation via video in addition to audio<br>➢ Prompts when BP exceeds an upper threshold or is below a lower threshold | ➢ Have option to note whether drug delivery was scheduled or Unscheduled.<br>➢ The system should track patients who have missed their deliveries |
|  | **Information to be collected on HIT** | ➢ Upon enrollment:<br>o Participant’s current prescription and the next refill date<br>o Encounter forms for peer delivery visits | ➢ Upon enrollment:<br>o Caregiver name, patient and caregiver contact information address<br>o Date patient started taking hypertension meds<br>o Name of peer assigned<br>o Clinic visit due date<br>➢ During delivery:<br>o BP reading and pill count<br>o Complaints/side effects<br>o Peer observation<br>o Any referrals made and to whom<br>o Provider details<br>o Refill due date |  |

#### Domain 1: Peer recruitment, training, and monitoring

Participants emphasized the importance of rigorous recruitment and adequate preparation of peers to support effective delivery of the PT4A implementation strategy. This led to consideration of which competencies should be prioritized during training. Proposed recruitment strategies included advertising positions through health facilities and community leadership structures, as well as clearly communicating eligibility criteria. Suggested criteria included completion of secondary education, age above 40 years, living with hypertension, and residence within the local community. Community health workers were also considered potential candidates because of their familiarity with patients and local health systems. Training would cover peer roles and responsibilities, blood pressure measurement, confidentiality, psychosocial support, HIT use, ethics and professionalism, referral procedures, and the medication delivery SOP, with refresher training provided as needed. Peer performance would be monitored using feedback from patients, clinicians, pharmacists, and community stakeholders, alongside indicators such as delivery accuracy and timeliness and participant enrollment and retention.

#### Domain 2: HIT

Participants identified the need for a reliable, user-friendly system that could support peers during medication delivery, particularly in settings with limited internet connectivity. Key requirements included timely medication refill reminders, easy data entry and retrieval, audio and video teleconsultation, and alerts for blood pressure readings outside recommended thresholds. In response, the team proposed an easy-to-navigate tablet-based system with offline data entry and synchronization once internet connectivity became available. The system would generate refill reminders approximately seven days before medications were due and alerts for blood pressure readings above or below predefined thresholds. It would also capture blood pressure readings, pill counts, medication information, patient complaints or side effects, peer observations, referrals, and delivery information, while support stock monitoring and tracking of refill and clinic appointment dates.

#### Domain 3: Medication delivery

Participants emphasized the need for a safe, reliable, and patient-centered process that ensured convenience, appropriate medication handling, and accurate and timely delivery. These considerations informed the development of a detailed SOP to standardize the delivery process. Peers and patients would communicate in advance regarding the delivery location and timing, and payment would be confirmed before medications were dispensed. Medication safety would be supported through appropriate packaging and storage. Patient feedback would be used to assess whether the correct medications were delivered on time and the quality of their interaction with the peer. Proof of delivery would be documented by the peer in the HIT system at the time of the delivery encounter and confirmed through the patient’s signed acknowledgment.

#### Domain 4: Medication payment

Participants identified the need for a secure, traceable, and convenient cashless payment process with minimal additional transaction costs. They also emphasized the importance of explaining payment procedures during informed consent so that participants understood the requirements before medication delivery began. The proposed approach was to use mobile payments exclusively, with payments made directly to hospital pharmacy till numbers to improve accountability and reduce transaction charges. Family support and participation in economic empowerment initiatives, such as table banking or farming groups, were suggested as potential mechanisms for patients who experienced difficulty paying for medications.

#### Domain 5: Stakeholder engagement

Participants emphasized early and continuous involvement of local authorities, communities, health facilities, county leadership, and relevant departments to strengthen acceptance and implementation of the intervention. Sustainability beyond the study period was also identified as an important consideration. The proposed stakeholders included county health leadership, health facility in-charges, county public health officers, clinicians, community health workers, chiefs, village elders, and representatives from similar AMPATH programs. Engagement meetings would introduce PT4A, clarify stakeholder roles, and establish mechanisms for continued involvement through meetings, visits, and telephone communication. For longer-term sustainability, the design team proposed engaging the Ministry of Health and county leadership regarding potential adoption of the intervention, including peer integration, medication supply arrangements, and continuation of the technology platform, while linking the intervention with existing community-based health programs.

### Acceptability and appropriateness testing

Among the healthcare workers (n = 18), the average age was 36.4 years, and the majority (56%) were female. The median number of years of clinical experience was six years, with a range of one to 25 years. Patients (n = 15) had an average age of 64 years, with the majority being female (53%). Ten of the patients had been diagnosed with hypertension for at least two years. Peers (n = 6) had an average age of 43.8 years, with the majority being male (67%). Peers’ median duration of experience was three years, with a range of two to 18 years.

**Figure 3** shows the distribution of total acceptability and appropriateness scores among healthcare workers, patients with hypertension, and peers. While average acceptability and appropriateness scores were relatively high in all three groups, both acceptability and appropriateness average scores were higher in patients and peers as compared to healthcare workers. Acceptability scores were highest among patients with hypertension (mean score of 19.3) and peers (mean score of 19.2). Healthcare workers had a lower mean acceptability score of 17.7. Appropriateness scores were highest amongst patients with hypertension and peers (mean score of 19.2 for both groups), while healthcare workers had a mean score of 17.3.

**Figure 3.**
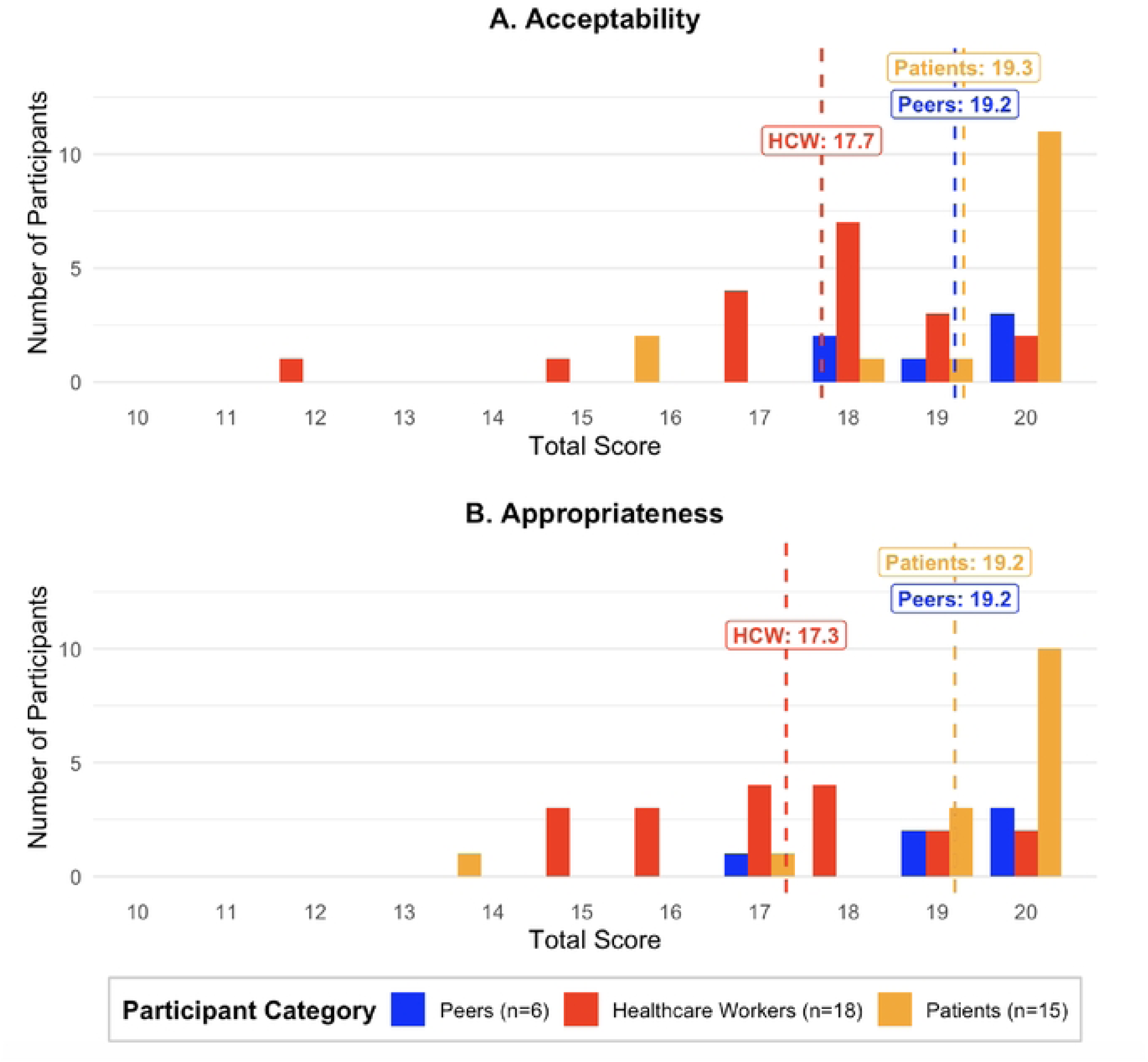
Frequency Distribution of Total Acceptability and Appropriateness Scores by Participant Category. Bar charts show the frequency distribution of total scores for acceptability (A) and appropriateness (B) of the PT4A implementation strategy. Peers (n = 6) are depicted in blue, healthcare workers (n = 18) are depicted in orange, and patients (n = 15) are depicted in red. The mean score for each participant group is indicated by the dashed line using the color corresponding to each participant category. *\*HCW = Healthcare Worker*

### Design meeting 5

In meeting 5, additional recommendations were generated to improve the acceptability and appropriateness of the implementation strategy for patients, peers, and healthcare workers. To strengthen peer support and oversight, the design team recommended that peers’ scope of work be clearly defined during training and shared this with clinicians to support monitoring and evaluation. The HIT tool was also refined to support oversight by enabling clinicians and the research team to monitor peer activities, track missed deliveries.

The team also recommended issuing identification badges to peers, clearly indicating their role as hypertension peers, to improve recognition and reduce stigma associated with home medication delivery. Additional recommendations included establishing clear procedures for medication delivery and developing a protocol to minimize loss to follow-up, including re-engaging patients through caregiver contact or home visits and specifying the number of re-contact attempts before classifying a patient as lost to follow-up.

The team also proposed more details on medication packaging, supply coordination, and healthcare worker engagement. Recommendations on medication packaging included: detailed receipts that would indicate the source of medications, medications dispensed, dispenser identity, and amount paid all to support transparency and accountability. Regular communication with pharmacists was also recommended to support consistent medication availability. To improve healthcare worker acceptability, the team recommended briefing clinicians on the study, patient eligibility criteria, peer qualifications, and peer training so that they could remain engaged throughout implementation. Non-monetary incentives, such as special training sessions, were also recommended to enhance clinician engagement.

## DISCUSSION

The PT4A strategy was developed in order to address significant barriers to hypertension medication adherence in western Kenya, specifically related to lack of access to and possession of medications, transportation-related challenges, and unmet psychosocial needs.^2,28^ Using a design team comprised of multiple different stakeholders, the PT4A strategy underwent refinement of operational details as well as appropriateness and acceptability testing, ultimately resulting in a prototype ready for pilot testing and implementation. The final prototype included the core components of the strategy from its inception--peer coaching, community-based medication delivery, and HIT--with additional refinements such as training requirements for peers, HIT usability improvements, and stipulations on financing and payment processes, to increase the likelihood of the strategy’s implementation success in the community. Overall, appropriateness and acceptability testing revealed that a majority of participants found that the revised implementation strategy was both appropriate and acceptable.

The HCD approach has proven useful in a number of settings to address several different healthcare delivery challenges and issues while increasing community and stakeholder participation in the design and implementation process. The pre-implementation and implementation “pipeline” encompass many different elements and stages, and HCD has been utilized at different stages of this process. Some studies have employed HCD at early time points, engaging stakeholders sometimes during the earliest stages of the design process in order to “co-design” the intervention.^12,30,31^ In contrast, other studies have involved HCD at later time points, often using stakeholder feedback to refine existing interventions.^32,33^

The optimal timing and duration of HCD in implementation research remains an area of debate and diversity of opinion. While there is consensus regarding the importance of a participatory approach and stakeholder involvement, there can be challenges for both stakeholders and investigators. Our team’s previous work in western Kenya has highlighted that there can sometimes be tension between implementation researchers and community stakeholders, especially when the “scaffolding” or initial specifications of an implementation strategy are “too fully” developed at the time that the HCD process is initiated.^10^ Our current approach, summarized here, involved clear communication throughout the design process between study investigators and design team members about the role of HCD, in this instance, to guide the development of operational details for the PT4A implementation strategy. This transparent communication helped to mitigate any potential tension and gave stakeholders a defined purpose in the process. As described above, our HCD process yielded valuable operational details regarding peer legitimacy, medication financing, and HIT that would have otherwise potentially gone un- or under-addressed without stakeholder perspectives. We believe that bringing together a diversity of stakeholders allowed for the entire group to feel activated to enact change collectively.^32^

Interestingly, however, despite the overall general success of the HCD process in supporting a diverse set of stakeholders to operationalize the PT4A strategy, healthcare workers gave markedly lower scores for acceptability and appropriateness compared to peers and patients. Reasons for this difference in acceptability and appropriateness may be attributed to the perception held by clinicians and healthcare workers that utilization of peers could be risky given the relative lack of extensive medical training that peers have compared to clinicians and other healthcare workers. To allay these concerns, several modifications were made in order to make the strategy more acceptable to clinicians by bolstering peer training and recruitment requirements, as described above. The differential perceptions across stakeholder groups thus did influence our team’s response to be more accommodating to the specific concerns of the different stakeholder groups. We anticipate that this approach will help to ensure that the implemented strategy maximizes buy-in by all stakeholder groups.

HCD as a form of stakeholder engagement is not without its critics, and we recognize that HCD is one among many potential participatory approaches that aim to secure stakeholder engagement and involvement. Alternative methods such as community-based participatory research, Delphi Method, and RAND/UCLA Appropriateness Method have been used in the past with success.^34–36^ Each approach has its own specific benefits and challenges; ideally, investigators and community stakeholders together can discuss and choose which approach is most appropriate for each particular situation, in order to optimize innovation, effectiveness, and equity in the context of resource limitations and other structural challenges.

In our situation specifically, we had scaffolding and a framework for our PT4A implementation strategy that we wanted to operationalize and implement in a specific region of western Kenya. When choosing among different participatory approaches, we prioritized stakeholder engagement in the design and refinement process of the PT4A strategy, with the eventual hope that we would foster stakeholder engagement for long-term success. We acknowledge, however, that the HCD process might not foster sustained engagement from stakeholders, potentially due to the duration and demand of long design phases and cycles that have previously been reported to result in stakeholder attrition.^7,37^ Stakeholder attrition becomes a particularly significant concern when one considers that not all participants are able to allocate the same amount of time to the design process, leading to situations where design teams may ultimately become less representative of the community they are serving by effectively selecting for those who are able to afford to dedicate time to the study.^38^ Because of the potential for stakeholder attrition and our understanding of the importance of clear communication with community members, we opted for a shorter HCD duration with involvement at a later time point in the design/operationalization process, recognizing potential shortcomings as described above. While it was a demanding experience that required flexibility and cooperation amongst a diversity of participants, we feel that our HCD approach ultimately was able to strike a balance between involving stakeholders to generate critical elements of the implementation strategy while not unduly burdening them.

We acknowledge the following limitations of our study. First, we recognize that while we did have a diverse group of stakeholders involved, not all potential stakeholder groups were represented. Future efforts could incorporate patient family members or caregivers who could provide input and insight regarding the impact of PT4A on patients and families. In addition, health facility-level administrators involved in scheduling appointments and staff assignments could also be included to provide the perspective of facility-level logistics and implications. Second, as we have discussed above, our choice of HCD is not the only participatory approach that could have been selected, and we recognize that other approaches could have their own benefits and drawbacks. Finally, our approach and the final PT4A implementation strategy might not be fully generalizable to other populations and other settings. However, we feel that the considerations raised by the design team, the operational details that were incorporated, and the final PT4A strategy can be relevant for populations and communities worldwide that are facing similar challenges of fragmented health systems, poverty, and transportation difficulties.

## CONCLUSION

We successfully used an HCD process to operationalize an implementation strategy to improve medication possession and adherence for hypertension management in western Kenya. The HCD process yielded valuable feedback from stakeholders, strengthening the implementation strategy and increasing its likelihood of long-term success, while also strengthening relationships between the research team and community stakeholders that can increase engagement and trust. We anticipate that future program planners, implementers, and implementation science researchers can build off our current work to continue engaging in successful participatory approaches to design and implement stronger, effective interventions to improve population health worldwide.

## Data Availability

The minimal data set will be made available on the NYU Langone Health Sciences Library Data Catalog. https://datacatalog.med.nyu.edu/

